# Fragile X Syndrome Carrier Screening Using a Nanopore Sequencing Assay

**DOI:** 10.1101/2024.07.23.24310865

**Authors:** Zhongmin Xia, Qiuxiao Deng, Ping Hu, Chunliu Gao, Yu Jiang, Yulin Zhou, Qiwei Guo

**Affiliations:** Department of Medical Genetics, Women and Children’s Hospital, School of Medicine, Xiamen University, Xiamen, Fujiang, China; School of Medicine, Xiamen University, Xiamen, Fujian, China; Department of Prenatal Diagnosis, Women’s Hospital of Nanjing Medical University, Nanjing Women and Children’s Healthcare Hospital, Nanjing, Jiangsu, China; School of Public Health, Xiamen University, Xiamen, Fujian, China; Molecular Diagnostic Laboratory for Precision Medicine, Xiang’an Hospital of Xiamen University, School of Medicine, Xiamen University, Xiamen, Fujian, China

**Keywords:** AGG interruption, carrier screening, CGG repeat, fragile X syndrome, nanopore sequencing

## Abstract

**BACKGROUND:** Fragile X syndrome (FXS) is the leading cause of monogenic autism spectrum disorder and inherited intellectual disabilities. Although the value of population-based FXS carrier screening has been acknowledged, appropriate screening methods are urgently required to establish and implement screening programs.

**METHODS:** We developed a nanopore sequencing-based assay that includes data analysis software to identify FXS carriers. Reference and clinical samples were used to evaluate the performance of our nanopore sequencing assay. Triplet-primed PCR and PacBio long-read sequencing were used for comparisons.

**RESULTS:** Nanopore sequencing identified reference carrier samples with a full range of premutation alleles in single-, 10-, and 100-plex assays, and identified AGG interruptions in an allele-specific manner. Nanopore sequencing revealed no size preference for amplicons containing different length CGG repeat regions. Finally, nanopore sequencing successfully identified three carriers among ten clinical samples for preliminary clinical validation. The observed variation in CGG repeat region size resulted from the base-calling process of nanopore sequencing.

**CONCLUSIONS:** Our nanopore sequencing assay is rapid, high-capacity, inexpensive, and easy to perform, thus providing a promising tool and paving the way for population-based FXS carrier screening.

## Introduction

Fragile X syndrome (FXS) (#MIM300624) is the leading cause of monogenic autism spectrum disorders and inherited intellectual disability, affecting approximately 1/7,000 females and 1/4,000 males worldwide (1, 2). The causative gene of FXS is fragile X messenger ribonucleoprotein 1 (*FMR1*), an X-linked dominant gene with full penetrance in all males and many females that plays a fundamental role in synapse formation and normal dendrite development (3, 4). *FMR1* can be categorized into four allelic forms based on the number of CGG trinucleotide repeats in its 5’ untranslated region: (a) normal alleles contain ~5 to ~44 repeats and are stable in meiosis or mitosis; (b) intermediate alleles contain ~45 to ~54 repeats and during intergenerational transmission, their repeat number can change slightly; (c) premutation alleles contain ~55 to ~200 repeats and can expand to full mutation during maternal transmission to offspring; premutation alleles are associated with risk of fragile X-associated primary ovarian insufficiency, fragile X-associated tremor/ataxia syndrome, and fragile X-associated neuropsychiatric disorders; (d) full mutation alleles have more than 200 repeats that frequently accompany hypermethylation in adjacent CpG islands and in the repeat region itself, which silences transcription and causes most cases of FXS (4). Asymptomatic women with pre-mutation or full mutation alleles are thus termed FXS carriers because their offspring are at risk of FXS (5). The prevalence of FXS carriers varies among different populations, ranging from approximately 1/149 in Israel to approximately 1/581 in East Asia (6–9). In many cases, CGG repeats are interrupted by one or more AGG trinucleotides (i.e., AGG interruptions), which can prevent strand slippage during replication, thus functioning as a protective factor that decreases the risk of intergenerational CGG expansion (9). Evaluation of AGG interruptions among CGG repeats is thus essential for genetic counseling, especially for FXS carriers (4).

Owing to the severe morbidity of FXS, carrier screening, and subsequent prenatal diagnosis are still warranted for this disease until the development of effective treatments (4, 10). Although all major ethnic groups and races appear to be susceptible to FXS, and FXS carrier prevalence is high, whether FXS carrier screening should be offered to the general population has long been a subject of debate (5, 9, 11–13). We previously showed that population-based carrier screening is the dominant strategy for FXS intervention in East Asia (9). Recently, population-based pan-ethnic FXS carrier screening was officially endorsed by the American College of Medical Genetics and Genomics (10). Clinical interest has shifted from “whether to perform” to “how to perform” population-based FXS carrier screening. An assay to capably address this issue should be able to examine a large number of female samples with high accuracy, low cost, short turnaround time, and ease of performance.

FXS genetic testing was performed using Southern blotting, a labor-intensive method that allows estimation of CGG expansion size but cannot accurately quantitate CGG repeat number and AGG interruption patterns (4). These drawbacks have been overcome with the triplet-primed PCR (TP-PCR) assay, which has become the mainstay method in clinical use, with several kits, including AmplideX^TM^ *FMR1* PCR Kit (Asuragen) and Molecular Fragile X PCR Kit (Biofast) commercially available (4, 14). Typing AGG interruptions at the allelic level in female samples with TP-PCR remains challenging. Long-read sequencing-based assays for FXS genetic testing using Nanopore and PacBio sequencing have recently been developed (15–19). Theoretically, with the ability to directly sequence entire CGG repeats and/or the full-length *FMR1* gene, long-read sequencing-based assays enable quantitative evaluation of CGG repeat number, AGG interruptions, rare intragenic variants, and large deletions in a single allele, dramatically advancing genetic diagnosis of FXS (16). To date, the clinical utility of long-read sequencing-based assays for population-based screening of FXS carriers has not been demonstrated.

In this study, we developed a nanopore sequencing assay that includes data analysis software and demonstrated its clinical potential as a promising tool for population-based FXS carrier screening.

## Materials and Methods

### SAMPLES

Eight (P2, P3, P4, P5, N1, N2, N3, and N4) and two (NA06968 and NA20239) reference samples with known *FMR1* genotypes were obtained from the National Institutes for Food and Drug Control of China and the Coriell Institute, respectively (20, 21). P3, P4, NA06968, and NA20239 provide genomic DNA from FXS carriers with different extents of CGG expansion, and N1, N2, N3, and N4 provide genomic DNA from women with normal *FMR1* alleles. N1/P2 and N1/P5 mixtures represent genomic DNA from women carrying CGG repeats near the intermediate/premutation boundary (Supplemental Table 1).

The genomic DNA of ten women whose *FMR1* genotypes have been examined via clinical FXS genetic testing was obtained from the Department of Prenatal Diagnosis, Women’s Hospital of Nanjing Medical University. Signed informed consent was obtained from each participant to authorize the use of their genetic data for research purposes. This study was approved by the Research Ethics Committee of Xiamen University.

### AMPLIFICATION OF *FMR1* ALLELES

The CGG repeats and flanking sequences of *FMR1* were amplified using a T100 Thermal Cycler (Bio-Rad, Hercules, CA, USA). A 50-µL reaction containing 1× Expand Long Template buffer 2 (Roche Diagnostics, Mannhei, Germany), 3.75U Expand Long Template Enzyme mix (Roche Diagnostics, Mannhei, Germany), 0.5 mmol/L dNTPs (Takara, Kyoto, Japan), 2.2 mol/L Betaine (SIGMA-Aldrich, Saint Louis, Missouri, USA), 0.33 mmol/L of each forward and reverse primer (Sangon, Shanghai, China), and 50 ng DNA template. Primer information and amplification conditions are listed in Supplemental Tables 2 and 3, respectively. After amplification, PCR products were purified with a TIANquick Midi Purification Kit (TIANGEN, Beijing, China) and subsequently quantified with a Qubit^™^ dsDNA HS Assay Kit (Invitrogen, Carlsbad, CA, USA) on a Qubit^™^ fluorometer (ThermoFisher Scientific, Waltham, Massachusetts, USA) according to the respective manufacturers’ instructions.

### LIBRARY PREPARATION AND NANOPORE SEQUENCING

Library preparation and nanopore sequencing methods are described in detail in Supplemental Methods.

### DATA ANALYSIS SOFTWARE

Based on the previously described STRique software (15), we developed data analysis software to facilitate the identification of CGG repeat numbers and AGG interruptions for FXS carrier screening in our nanopore sequencing assay. The logic of software is described in detail in Suppmental Methods; the software code was licensed by Xiamen University and is available at https://github.com/guoqiwei-xmu/FXS-carrier-identifier.git.

### TP-PCR ASSAY

A CE-certified AmplideX^™^ FMR1 PCR Kit (Asuragen, Minneapolis, Minnesota, USA) and a Chinese Food and Drug Administration-approved Molecular Fragile X PCR Kit (Biofast, Xiamen, Fujian, China) were used as comparative methods. Respective assays were performed on a T100 Thermal Cycler (Bio-Rad, Hercules, CA, USA) and a 3500 DX Genetic Analyzer (Applied Biosystems, Waltham, Massachusetts, USA), according to the manufacturers’ instructions.

### CAFXS ASSAY

A long-read sequencing-based method termed the CAFXS assay, was used as a comparative method and performed by a commercial service (Berry Genomics, Beijing, China). The principle of CAFXS has been previously described (16).

## Results

### DESIGN OF NANOPORE SEQUENCING ASSAY FOR FXS CARRIER SCREENING

As illustrated in Fig. 1, our nanopore sequencing assay comprises three steps: pre-sequencing preparation, nanopore sequencing, and post-sequencing data analysis. During pre-sequencing preparation, CGG repeats and flanking sequences of each sample are amplified with a pair of primers containing a specific identifier sequence at each strand’s 5’ end. These primers allow the specific identifier sequence to be integrated into the amplicon of each sample to enable the analysis of multiple samples in a single sequencing assay. After purification and quantification, amplicons are pooled to achieve equal masses to prepare the sequencing library. After nanopore sequencing of the library, post-sequencing data analysis is performed using novel data analysis software. Briefly, the nucleotide sequence of each read is translated from a raw ionic current signal (base calling), and qualified reads that contain correct identifier, prefix, and suffix sequences, are retained for further analysis. Next, the CGG repeat numbers of qualified reads are evaluated based on the distance between prefix and suffix sequences. After, the presence and position of AGG interruptions are evaluated for each qualified read. For each sample, the distribution of reads with different CGG repeat numbers and AGG statuses is analyzed. Finally, the CGG repeat number and AGG status of a specific allele are determined based on the peaks generated from the aggregated reads with similar CGG repeat numbers and AGG patterns. When an allele with a CGG repeat number over the threshold (e.g., 54 or 55) is identified, an alarm is triggered to flag an FXS carrier.

**Figure 1.**
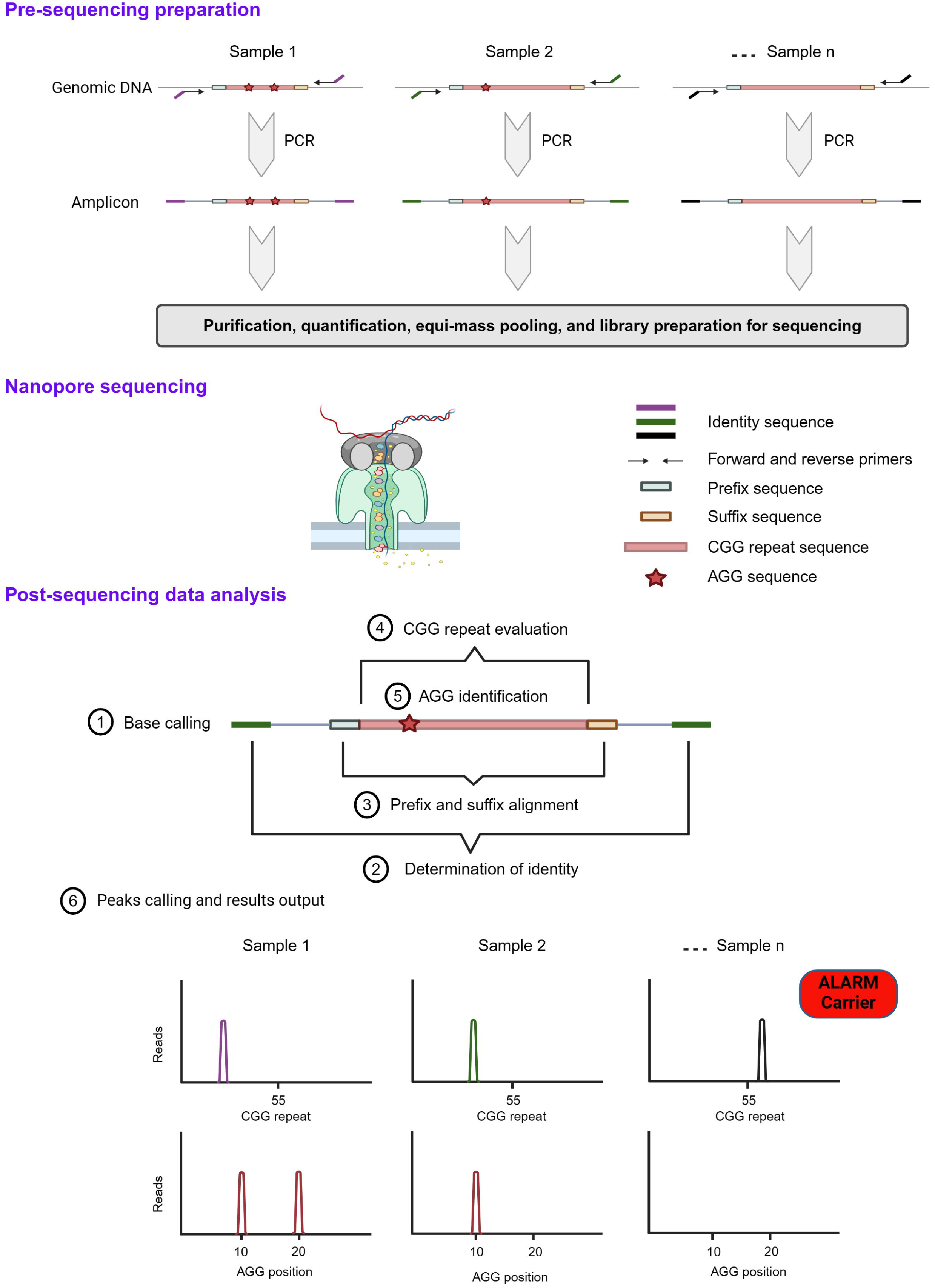
Flowchart of screen for fragile X syndrome carriers using our nanopore sequencing assay.

### VALIDATION OF NANOPORE SEQUENCING ASSAY BY EXAMINING FXS CARRIERS WITH VARIOUS CGG REPEAT NUMBERS AND AGG INTERRUPTION PATTERNS

We first validated the nanopore sequencing assay by examining FXS carriers with various numbers of CGG repeats and AGG interruption patterns in a single-plex manner. Reference samples were used as FXS carriers, and TP-PCR and CAFXS were used as methods for comparison. As shown in Fig. 2, nanopore sequencing identified a wide range of expanded alleles. AGG interruption patterns were identified in an allele-specific manner. A slight discordance between nanopore sequencing and comparative method results was observed in the reported CGG repeat numbers of expanded alleles (Table 1). Due to a one CGG repeat difference between methods, the N1/P5 sample was designated as a non-carrier by the nanopore sequencing assay but a carrier by comparative methods.

**Figure 2.**
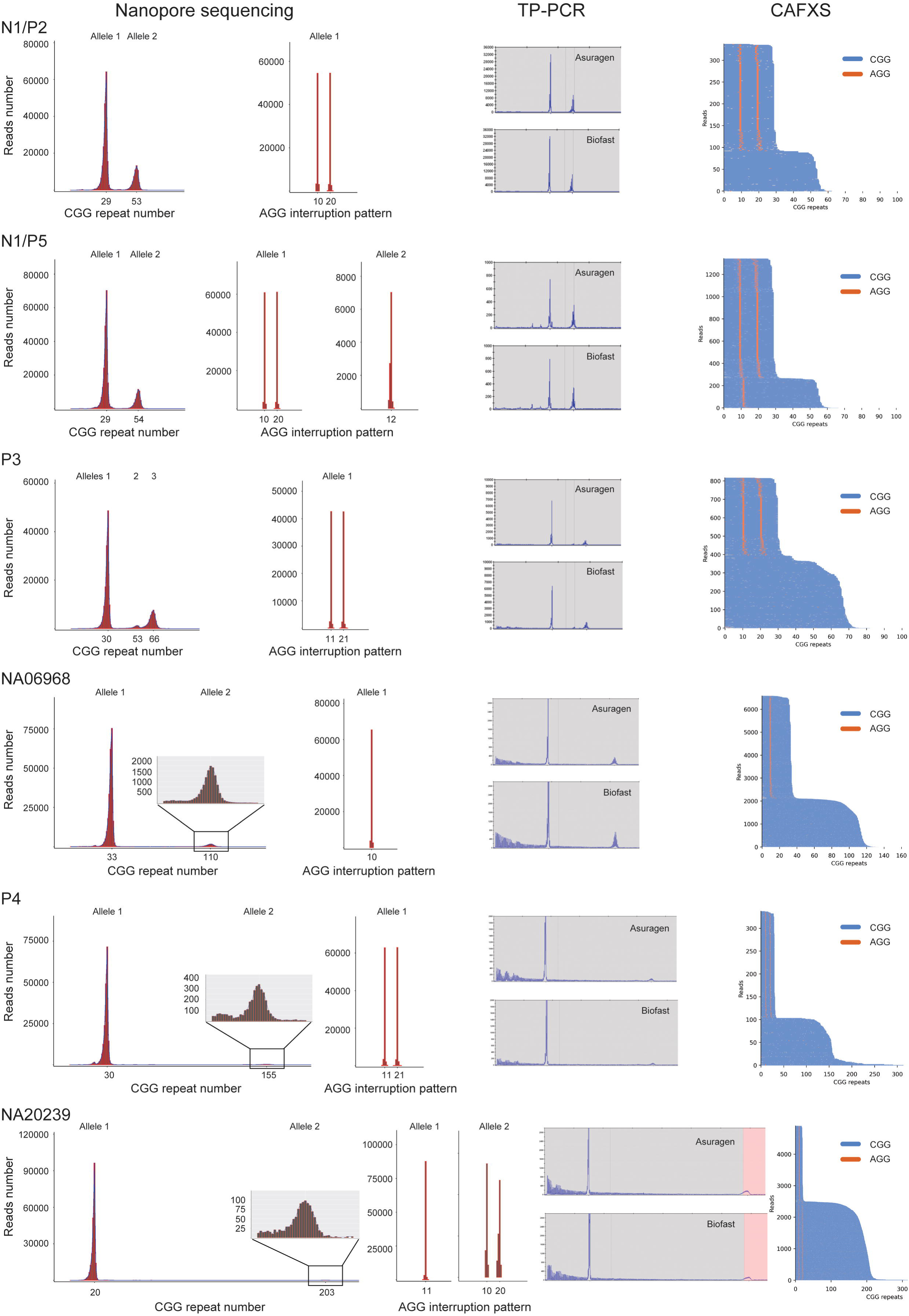
Comparison of nanopore sequencing, TP-PCR, and CAFXS for detection of different FXS carrier reference samples.

**Table 1.**
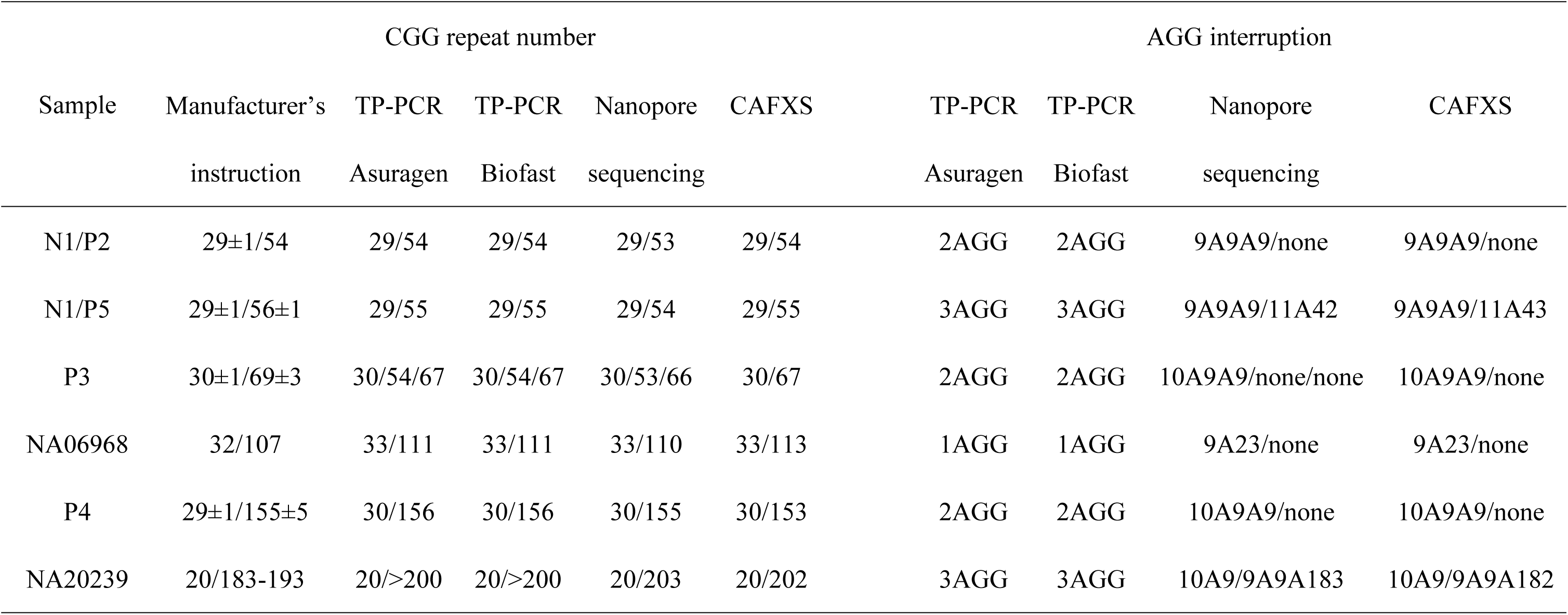
Comparison of nanopore sequencing, TP-PCR, and CAFXS for the detection of different FXS carrier reference samples.

### DETECTION OF FXS CARRIERS FROM WILDTYPE BACKGROUNDS WITH THE MULTIPLEX NANOPORE SEQUENCING ASSAY

We next evaluated whether the nanopore sequencing assay could examine multiple samples simultaneously and distinguish FXS carriers from wild-type backgrounds, as shown in Fig. 1. Ten pairs of primers labeled with different identifier sequences were designed, validated, and used to amplify ten respective samples (Fig. 3A and Supplemental Table 2). As illustrated in Fig. 1, amplicons were pooled to achieve equal masses for library preparation and nanopore sequencing. Nanopore sequencing successfully detected a wide range of expanded alleles in the 10-plex assay (Fig. 3B-G).

**Figure 3.**
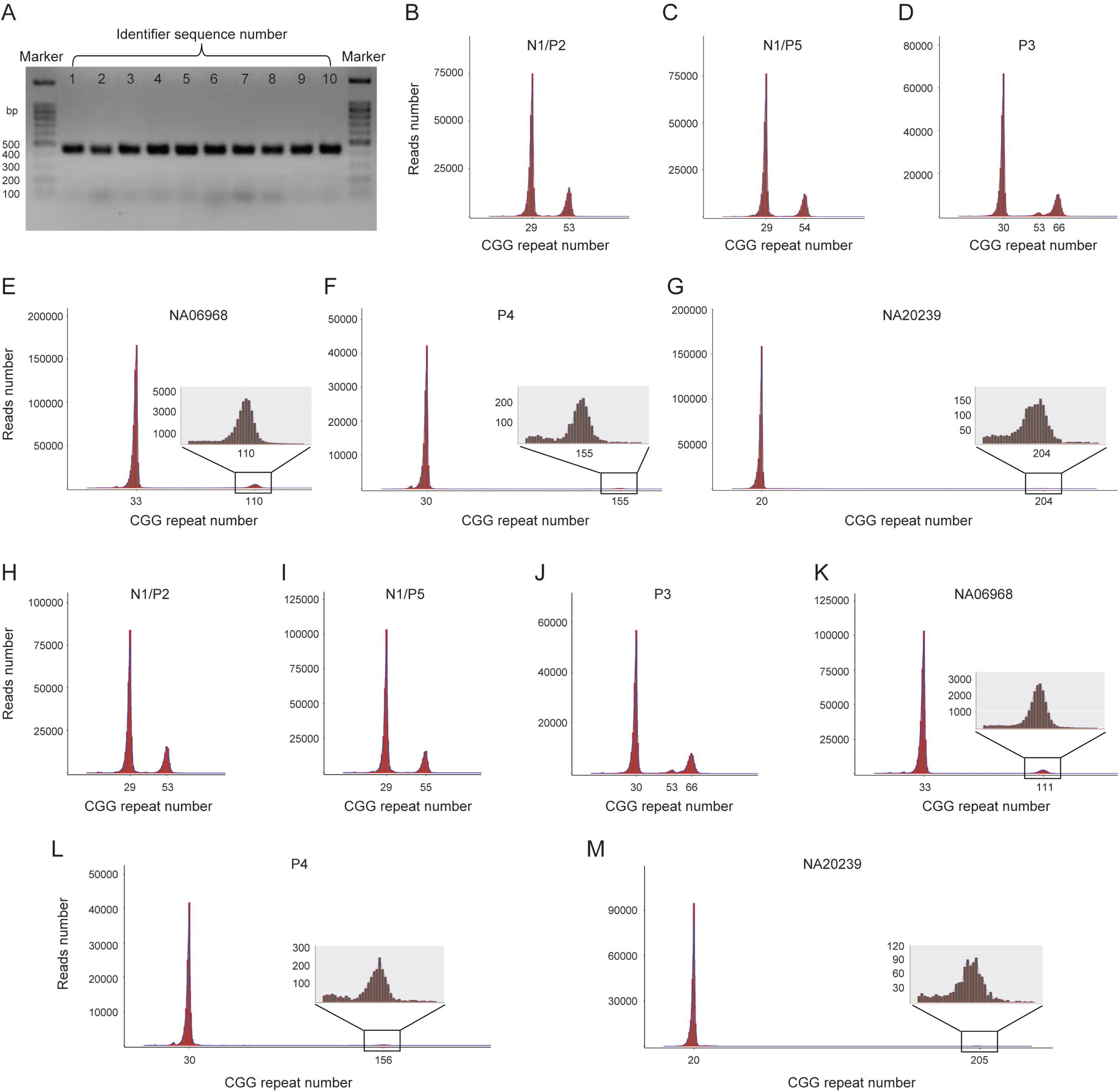
Detection of FXS carriers within wildtype backgrounds with multiplex nanopore sequencing assay. (A) Efficacy of ten pairs of primers labeled with different identifier sequences was validated by amplification of wildtype samples. (B-G) CGG repeat numbers associated with reference samples in the 10-plex nanopore sequencing assay. (H-M) CGG repeat numbers associated with reference samples in the 100-plex nanopore sequencing assay.

We assessed whether the nanopore sequencing assay could simultaneously examine more samples (e.g., 100 samples). Instead of using 100 identifier sequences to identify 100 different samples, we mimicked the scenario by decreasing the pooling proportion of the six target amplicons to 6% (1% each), while increasing the pooling proportion of the four wild-type amplicons to 94% (23.5% each). Similar to the results of the 10-plex assay, the 100-plex assay successfully detected all expanded alleles within wild-type backgrounds (Fig. 3H-M).

### INACCURATE BASE CALLING ATTRIBUTED TO VARIATIONS IN CGG REPEAT QUANTIFICATION

Although the nanopore sequencing assay was able to detect various FXS carriers, the reported CGG repeat numbers of expanded alleles differed slightly from those of the comparative methods (Table 1). Moreover, for a specific reference sample, the reported CGG repeat numbers of the expanded alleles were slightly discordant among the different nanopore sequencing assays (Fig. 4A). To investigate the potential causes of these variations, we manually examined the sequences of 20 random reads with fewer CGG repeats than those expected in the nanopore sequencing assay. As shown in Fig. 4B, sequence variants due to inaccurate base calling were distributed in these reads, including prefix, suffix, and CGG repeat sequences, resulting in the miscalculation of CGG repeat numbers in the subsequent data analysis processes. We analyzed the Fastq data (i.e., data after base calling) for N1/P2 and N1/P5 reference samples derived from the CAFXS assay using our data analysis software, and the reported CGG repeat numbers of these reference samples were concordant between our data analysis pipeline and the CAFXS assay (Supplemental Fig. 1) confirming the accuracy of post-base-calling processes using our data analysis software. Lastly, we reanalyzed N1/P2 and N1/P5 reference samples based on sequencing data derived from the sense and antisense strands, respectively, and confirmed that there was no strand bias in CGG repeat quantification (Fig. 4C). Collectively, the observed variations in CGG repeat quantification are thus mainly attributed to inaccurate base-calling.

**Figure 4.**
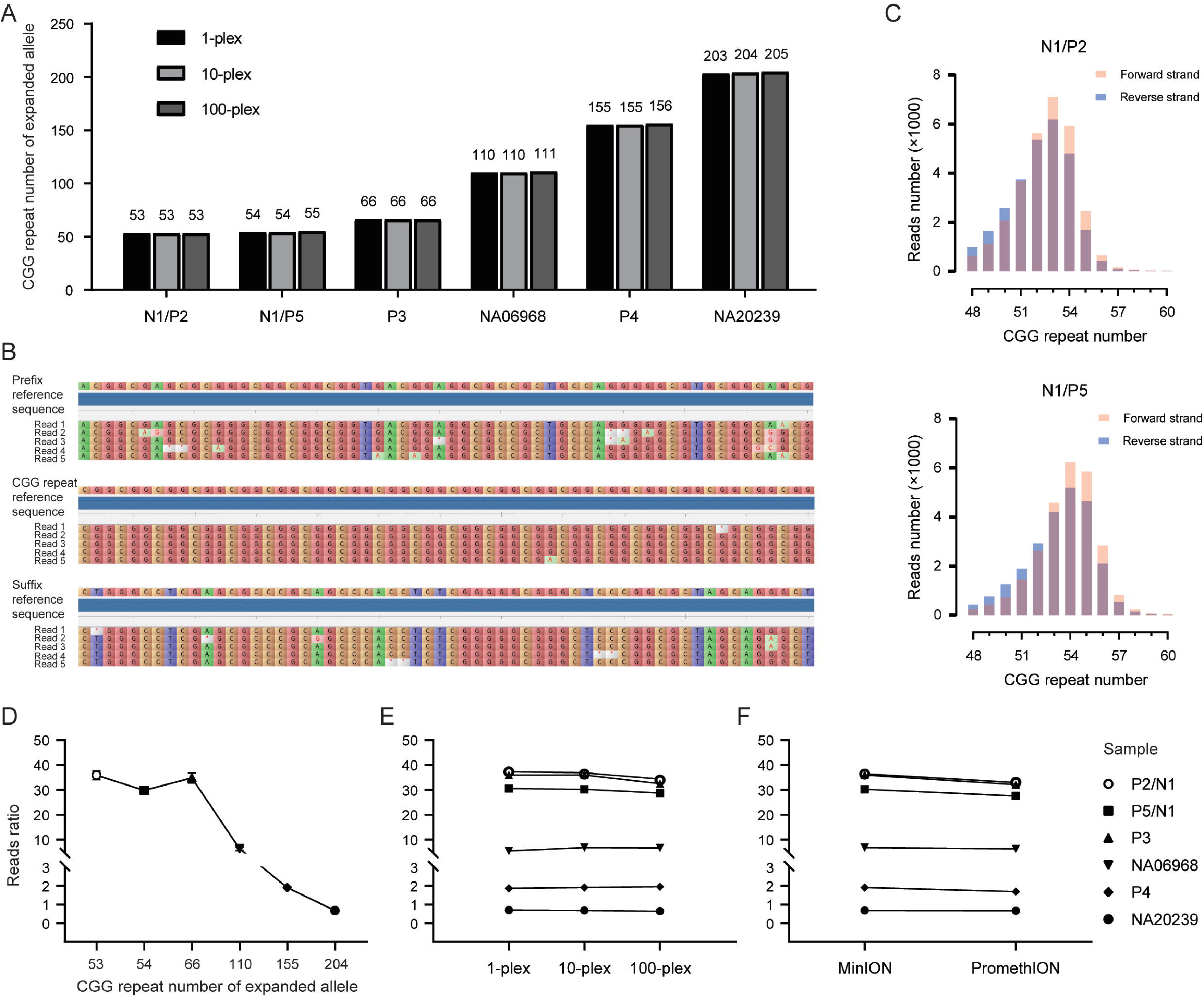
Evaluation of length variation and length preference of the nanopore sequencing assay. (A) Slight variations were observed in reported CGG repeat numbers for expanded alleles in reference samples among single-plex, 10-plex, and 100-plex nanopore sequencing assays. (B) Representative reads with sequence variants due to inaccurate base calling. (C) Reported CGG repeat numbers of expanded alleles were identical based on reads derived from forward and reverse strands, respectively. (D) Read ratios of expanded alleles versus their wild-type counterparts tend to decrease with increasing allelic expansion. Error bars indicate standard deviation among triplicate samples. (E) Reads ratios of expanded alleles versus their wildtype counterparts were similar among single-plex,10-plex, and 100-plex nanopore sequencing assays. (F) Reads ratios of expanded alleles versus their wildtype counterparts were similar between MinION and PromethION platforms whose respective flow cells had 1190 and 8657 active nanopores.

### NANOPORE SEQUENCING DEMONSTRATES NO SIZE PREFERENCES FOR AMPLICONS WITH DIFFERENT CGG REPEATS

Although the nanopore sequencing assay could detect FXS carriers with a wide range of expanded alleles, we noticed that for a specific carrier, the read ratios of expanded alleles versus their wild-type counterparts tended to decrease with increasing allelic expansion (Fig. 4D). We thus evaluated whether the short amplicons of the wild-type alleles more readily passed through the nanopore than the longer amplicons of the expanded alleles. Two libraries derived from reference samples, N1 (29 CGG repeats) and P5 (55 CGG repeats) were pooled in equimolar amounts and sequenced. The read ratio of P5 to N1 was approximately 1.06, suggesting that there was no size preference for amplicons with different CGG repeats (Supplemental Fig. 2). Moreover, we compared the read ratios of the expanded alleles with their wild-type counterparts using single-plex, 10-plex, and 100-plex nanopore sequencing assays. As shown in Fig. 4E, the read ratios of each sample were similar among the three assays, suggesting that in the presence of different wild-type backgrounds, large amounts of short amplicons did not skew the target size-indiscriminative characteristics of nanopore sequencing. Finally, we evaluated the same samples on different nanopore sequencing platforms, namely MinION and PromethION, which had 1190 and 8657 active nanopores on their flow cells at the beginning of sequencing, respectively. As shown in Fig. 4F, the read ratios were similar between the two platforms, suggesting that the target-size indiscriminatory characteristics of nanopore sequencing were not influenced by the number of active nanopores in the flow cells.

### PRELIMINARY VALIDATION OF CLINICAL UTILITY OF NANOPORE SEQUENCING ASSAY FOR THE FXS CARRIER SCREENING

To validate the clinical utility of the nanopore sequencing assay for FXS carrier screening, we analyzed 10 female clinical samples using a 10-plex nanopore sequencing assay. Considering the variations in CGG repeat quantification, when an allele with ≥ 54 CGG repeats was identified, an alert would be triggered designating a carrier. These samples were analyzed in parallel with CAFXS assay data. As shown in Table 2 and Supplemental Fig. 3, three carriers were identified using the nanopore sequencing assay, concordant with the results of the CAFXS assays.

**Table 2.** Evaluation of nanopore sequencing assay with clinical samples.

| Sample | CGG repeat number |  | AGG interruption |  |
| --- | --- | --- | --- | --- |
|  | Nanopore | CAFXS | Nanopore | CAFXS |
|  | sequencing |  | sequencing |  |
| 1* | 36/151 | 36/153 | 9A9A6A9/9A141 | 9A9A6A9/9A143 |
| 2 | 33/50 | 33/51 | 9A6A6A9/10A39 | 9A6A6A9/10A40 |
| 3 | 29/52 | 29/53 | 9A9A9/9A9A5A9A6A9 | 9A9A9/9A9A6A9A6A9 |
| 4* | 29/54 | 29/55 | 9A9A9/9A9A10A6A6A9 | 9A9A9/9A9A11A6A6A9 |
| 5* | 36/60 | 36/61 | 9A9A6A9/10A49 | 9A9A6A9/10A50 |
| 6 | 29/29 | 29/29 | 9A9A9/9A9A9 | 9A9A9/9A9A9 |
| 7 | 20/30 | 20/30 | 10A9/none | 10A9/none |
| 8 | 29/29 | 29/29 | 9A9A9/9A9A9 | 9A9A9/9A9A9 |
| 9 | 36/36 | 36/36 | 9A9A6A9/9A16A9 | 9A9A6A9/9A16A9 |
| 10 | 26/29 | 26/29 | 10A9A5/9A9A9 | 10A9A5/9A9A9 |

\*FXS carrier

## Discussion

Given that the value of population-based FXS carrier screening has been acknowledged (10), appropriate screening methods are urgently needed to implement such screening programs. A favorable testing method should be able to examine large-scale samples with high accuracy, low cost, short turnaround times, and ease of performance. Our nanopore sequencing assay would be one option that can meet these demands. First, we developed reliable and user-friendly software to facilitate sequencing data analysis and output results. Second, using reference and clinical samples, we demonstrated that the nanopore sequencing assay was able to identify the full range of premutation *FMR1* alleles, which account for the majority of FXS carriers, and the nanopore sequencing assay was able to reliably quantify CGG repeats and AGG interruptions, which are two important determinants for assessing the risk of full mutation expansion in carriers, thus facilitating genetic counseling for carrier screening. Third, using identifier sequences, we tested multiple samples in one assay, dramatically increasing test capacity and decreasing cost. In our 100-plex assay, the cost per sample was less than USD 10. Moreover, nanopore sequencing demonstrated no size preferences for amplicons with different CGG repeats, suggesting that the detection of expanded alleles will not be compromised by wild-type allele abundance as long as sufficient data are collected for each sample, while tens of thousands of identifier sequences are available in the form of different short combinations of nucleotides. In this regard, we could further increase sample capacity and decrease cost per sample by collecting more data using an advanced nanopore sequencing platform, such as PromethION. Fourth, the nanopore sequencing assay can be easily accomplished with a short turnaround time (for example, a 100-plex assay requires approximately two days) by a technician in a regular molecular diagnostics laboratory. Lastly, in comparison with TP-PCR and CAFXS, the equipment for nanopore sequencing assays is more available and portable, and thus could be more readily and widely adapted, particularly in underdeveloped regions of the world (22).

In comparison with NGS or PacBio-based long-read sequencing, nanopore sequencing has a relatively higher sequencing error rate in single reads owing to inaccuracies in ionic current signal-to-nucleotide sequence translation (i.e., the base calling process) (22). This innate error would cause one to several copy number variations during quantification of CGG repeats in our nanopore assays, which may cause misclassification of subjects whose CGG repeat numbers are near the boundary between intermediate and premutation alleles. Technical limitation-associated variations in CGG repeat sizing are common to most methods and are allowed to a certain extent in technical standards (4). The accuracy of CGG repeat sizing can be improved, rendering it not a major concern in the use of our nanopore sequencing assay. For example, base calling is educable with machine learning models, and its accuracy continuously increases with the accumulation of nanopore sequencing data (23–25). Even at the current stage, a reference sample, such as N1/P5, can be used as a quantitative control to monitor variation in base calling and direct the threshold setting for carrier identification.

In comparison with amplification-free methods (15, 18), amplification of *FMR1* alleles drastically increases the sequencing depth of the target region and decreases costs. However, owing to the high GC content of the target region and competitive effects of wild-type alleles, the amplification efficiencies of expanded alleles tend to decrease with increasing allelic expansion (4). In this study, carriers with a full range of premutation alleles were identified by our nanopore sequencing assay; however, carriers with full mutation alleles, which constitute approximately 3% of FXS carriers (9), could be missed by our assay. This issue can be partially resolved by further optimizing the amplification system through the optimization of reagents or reaction conditions. While the sensitivity of mosaicism detection has not been systematically examined because its clinical significance in carrier screening remains unclear, this sensitivity could be compared between nanopore sequencing and TP-PCR (~5-10%) (26) using the mosaicism detection results of reference sample P3.

In conclusion, we developed a nanopore sequencing-based assay to identify FXS carriers that includes data analysis software. This assay was demonstrated to be rapid, high-capacity, inexpensive, and easy to perform, thus providing a promising tool for population-based FXS carrier screening.

## Research Funding

This work was supported by the National Natural Science Foundation of China [grant numbers 82071662] and the Natural Science Foundation of Xiamen Municipality [grant numbers 3502Z20227141].

## Data Availability

All data produced in the present work are contained in the manuscript

## Nonstandard abbreviations

FXS: fragile X syndrome
TP-PCR: triplet-primed PCR

## Human gene

*FMR1*: fragile X messenger ribonucleoprotein 1

## Acknowledgement

We thank all the clinical participants, and I-Fan Chiu, Maoli Chen, Sihao Wu, Hanwei Wang, and Lingfeng Mao for their kind supports to this work.

## References

1. Mila M, Alvarez-Mora MI, Madrigal I, Rodriguez-Revenga L. Fragile x syndrome: An overview and update of the fmr1 gene. Clinical Genetics 2017;93:2:197–205 doi: 10.1111/cge.13075.

2. Lozano R, Azarang A, Wilaisakditipakorn T, Hagerman RJ. Fragile x syndrome: A review of clinical management. Intractable & Rare Diseases Research 2016;5:3:145–57 doi: 10.5582/irdr.2016.01048.

3. Pieretti M, Zhang F, Fu Y-H, Warren ST, Oostra BA, Caskey CT, et al. Absence of expression of the fmr-1 gene in fragile x syndrome. Cell 1991;66:4:817–22 doi: 10.1016/0092-8674(91)90125-i.

4. Spector E, Behlmann A, Kronquist K, Rose NC, Lyon E, Reddi HV. Laboratory testing for fragile x, 2021 revision: A technical standard of the american college of medical genetics and genomics (acmg). Genet Med 2021;23:5:799–812. Epub 20210401 doi: 10.1038/s41436-021-01115-y.

5. Committee opinion no. 691: Carrier screening for genetic conditions. Obstet Gynecol 2017;129:3:e41–e55. Epub 2017/02/23 doi: 10.1097/AOG.000000000000195200006250-201703000-00048 [pii].

6. Zlotogora J, Grotto I, Kaliner E, Gamzu R. The israeli national population program of genetic carrier screening for reproductive purposes. Genetics in Medicine 2016;18:2:203–6 doi: 10.1038/gim.2015.55.

7. Westemeyer M, Saucier J, Wallace J, Prins SA, Shetty A, Malhotra M, et al. Clinical experience with carrier screening in a general population: Support for a comprehensive pan-ethnic approach. Genetics in Medicine 2020;22:8:1320–8 doi: 10.1038/s41436-020-0807-4.

8. Archibald AD, Smith MJ, Burgess T, Scarff KL, Elliott J, Hunt CE, et al. Reproductive genetic carrier screening for cystic fibrosis, fragile x syndrome, and spinal muscular atrophy in australia: Outcomes of 12,000 tests. Genet Med 2018;20:5:513–23. Epub 20171026 doi: 10.1038/gim.2017.134.

9. Guo Q, Chang YY, Huang CH, Hsiao YS, Hsiao YC, Chiu IF, et al. Population-based carrier screening and prenatal diagnosis of fragile x syndrome in east asian populations. J Genet Genomics 2021;48:12:1104–10. Epub 20210607 doi: 10.1016/j.jgg.2021.04.012.

10. Gregg AR, Aarabi M, Klugman S, Leach NT, Bashford MT, Goldwaser T, et al. Screening for autosomal recessive and x-linked conditions during pregnancy and preconception: A practice resource of the american college of medical genetics and genomics (acmg). Genetics in Medicine 2021;23:10:1793–806 doi: 10.1038/s41436-021-01203-z.

11. Dimmock DP. Should we implement population screening for fragile x? Genetics in Medicine 2017;19:12:1295–9 doi: 10.1038/gim.2017.81.

12. Metcalfe SA, Delatycki MB, Cohen J, Archibald AD, Emery JD. Fragile x population carrier screening. Genetics in Medicine 2018;20:9:1091–2 doi: 10.1038/gim.2017.209.

13. Metcalfe SA, Martyn M, Ames A, Anderson V, Archibald AD, Carter R, et al. Informed decision making and psychosocial outcomes in pregnant and nonpregnant women offered population fragile x carrier screening. Genetics in Medicine 2017;19:12:1346–55 doi: 10.1038/gim.2017.67.

14. Juusola JS, Anderson P, Sabato F, Wilkinson DS, Pandya A, Ferreira-Gonzalez A. Performance evaluation of two methods using commercially available reagents for pcr-based detection of fmr1 mutation. J Mol Diagn 2012;14:5:476–86. Epub 20120702 doi: 10.1016/j.jmoldx.2012.03.005.

15. Giesselmann P, Brändl B, Raimondeau E, Bowen R, Rohrandt C, Tandon R, et al. Analysis of short tandem repeat expansions and their methylation state with nanopore sequencing. Nat Biotechnol 2019;37:12:1478–81. Epub 20191118 doi: 10.1038/s41587-019-0293-x.

16. Liang Q, Liu Y, Duan R, Meng W, Zhan J, Xia J, et al. Comprehensive analysis of fragile x syndrome: Full characterization of the fmr1 locus by long-read sequencing. Clin Chem 2022;68:12:1529–40. Epub 2022/09/29 doi: 10.1093/clinchem/hvac1546726663 [pii].

17. Ardui S, Race V, Zablotskaya A, Hestand MS, Van Esch H, Devriendt K, et al. Detecting agg interruptions in male and female fmr1 premutation carriers by single-molecule sequencing. Human Mutation 2017;38:3:324–31 doi: 10.1002/humu.23150.

18. Stevanovski I, Chintalaphani SR, Gamaarachchi H, Ferguson JM, Pineda SS, Scriba CK, et al. Comprehensive genetic diagnosis of tandem repeat expansion disorders with programmable targeted nanopore sequencing. Sci Adv 2022;8:9:eabm5386. Epub 20220304 doi: 10.1126/sciadv.abm5386.

19. Loomis EW, Eid JS, Peluso P, Yin J, Hickey L, Rank D, et al. Sequencing the unsequenceable: Expanded cgg-repeat alleles of the fragile x gene. Genome Research 2013;23:1:121–8 doi: 10.1101/gr.141705.112.

20. Amos Wilson J, Pratt VM, Phansalkar A, Muralidharan K, Highsmith WE, Beck JC, et al. Consensus characterization of 16 fmr1 reference materials: A consortium study. The Journal of Molecular Diagnostics 2008;10:1:2–12 doi: 10.2353/jmoldx.2008.070105.

21. Gao F, Huang W, You Y, Huang J, Zhao J, Xue J, et al. Development of chinese genetic reference panel for fragile x syndrome and its application to the screen of 10,000 chinese pregnant women and women planning pregnancy. Molecular Genetics & Genomic Medicine 2020;8:6 doi: 10.1002/mgg3.1236.

22. Ameur A, Kloosterman WP, Hestand MS. Single-molecule sequencing: Towards clinical applications. Trends in Biotechnology 2019;37:1:72–85 doi: 10.1016/j.tibtech.2018.07.013.

23. Singh G, Alser M, Denolf K, Firtina C, Khodamoradi A, Cavlak MB, et al. Rubicon: A framework for designing efficient deep learning-based genomic basecallers. Genome Biology 2024;25:1 doi: 10.1186/s13059-024-03181-2.

24. Amarasinghe SL, Su S, Dong X, Zappia L, Ritchie ME, Gouil Q. Opportunities and challenges in long-read sequencing data analysis. Genome Biology 2020;21:1 doi: 10.1186/s13059-020-1935-5.

25. Alser M, Lindegger J, Firtina C, Almadhoun N, Mao H, Singh G, et al. From molecules to genomic variations: Accelerating genome analysis via intelligent algorithms and architectures. Computational and Structural Biotechnology Journal 2022;20:4579–99 doi: 10.1016/j.csbj.2022.08.019.

26. Hantash FM, Goos DG, Tsao D, Quan F, Buller-Burckle A, Peng M, et al. Qualitative assessment of fmr1 (cgg)n triplet repeat status in normal, intermediate, premutation, full mutation, and mosaic carriers in both sexes: Implications for fragile x syndrome carrier and newborn screening. Genetics in Medicine 2010;12:3:162–73 doi: 10.1097/GIM.0b013e3181d0d40e.

