## Supplementary figures and images for "Fragile X Syndrome Carrier Screening Using a Nanopore Sequencing Assay"

### Supplemental Fig. 1

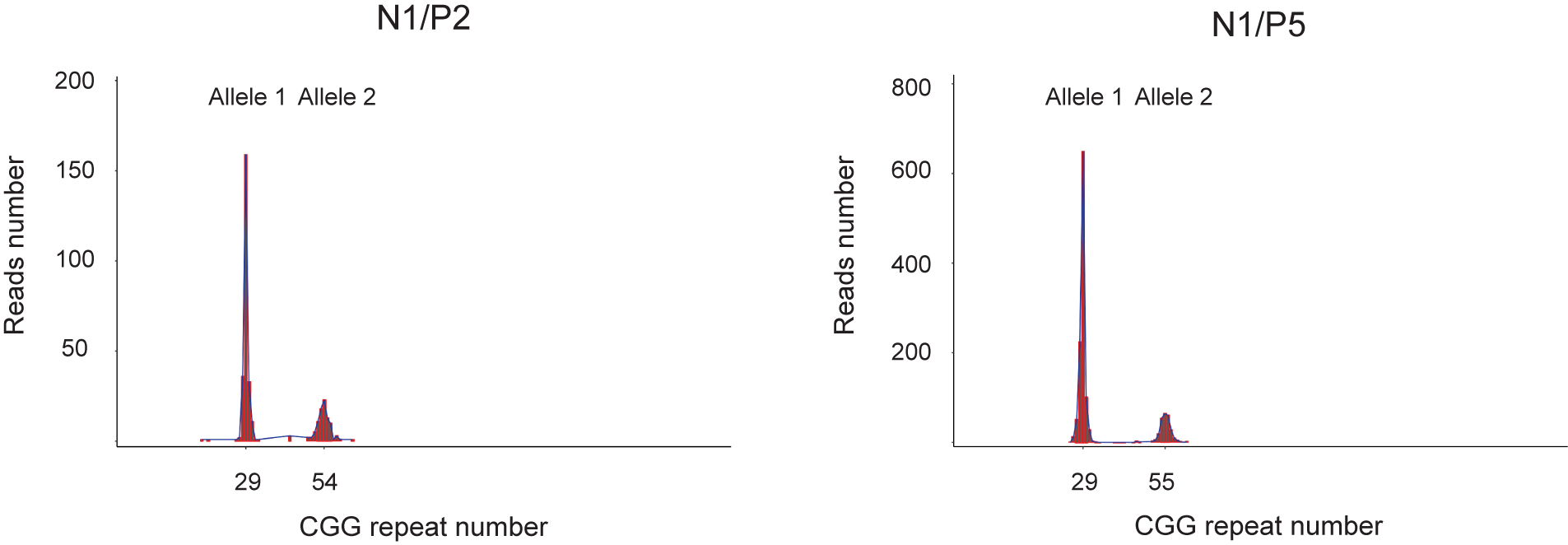

### Supplemental Fig. 2

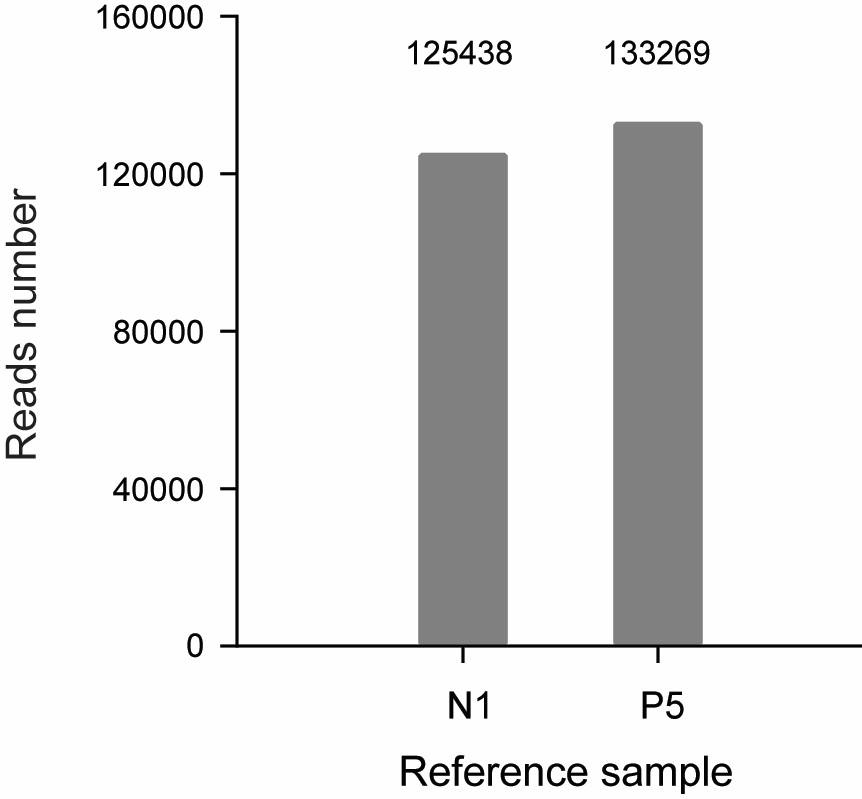

### Supplemental Fig. 3

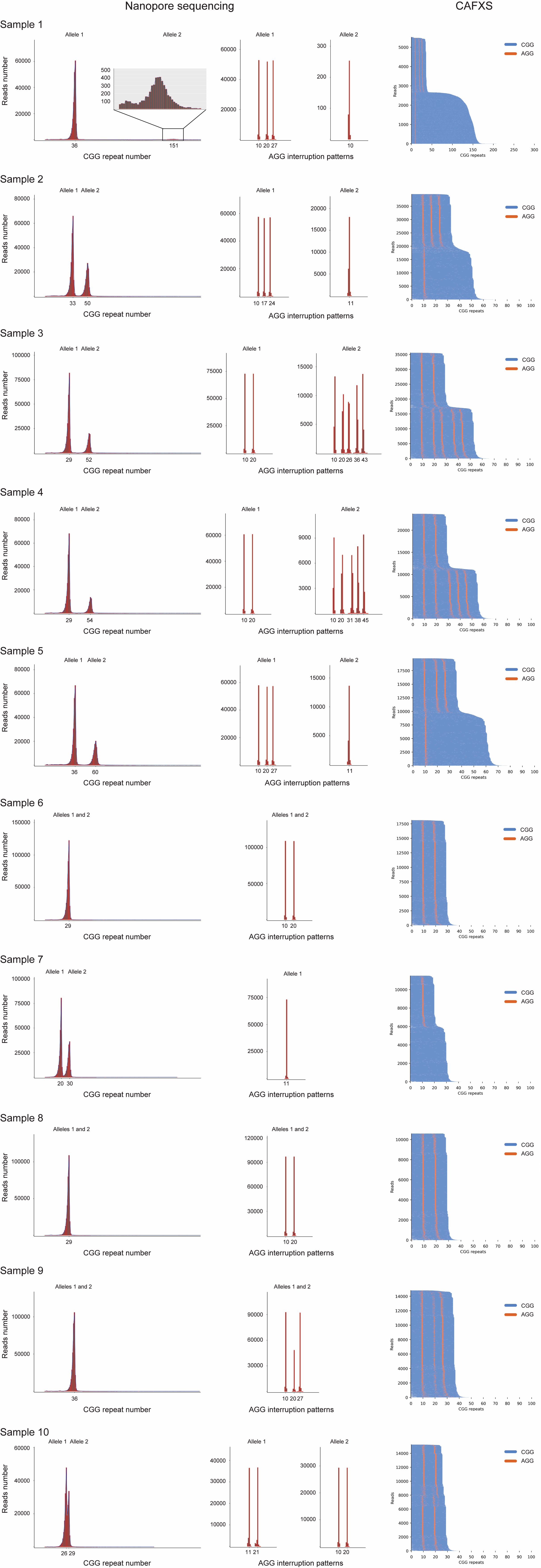
